# Molecular landscape of pelvic organ prolapse provides insights into disease etiology and clues towards putative novel treatments

**DOI:** 10.1101/2020.03.12.20034165

**Authors:** Kirsten B. Kluivers, Sabrina L. Lince, Alejandra M. Ruiz-Zapata, Rufus Cartwright, Manon H. Kerkhof, Joanna Widomska, Ward De Witte, Wilke M. Post, Jakub Pecanka, Lambertus A. Kiemeney, Sita H. Vermeulen, Jelle J. Goeman, Kristina Allen-Brady, Egbert Oosterwijk, Geert Poelmans

## Abstract

**Background:** Pelvic organ prolapse (POP) represents a major health care burden in women but its underlying pathophysiological mechanisms have not been elucidated.

**Objective:** To integrate the results from a large scale exome chip study with published genetic and expression data into a molecular landscape of POP.

**Design, setting, and participants:** The exome chip study was conducted in 526 women with POP and 960 healthy controls. To corroborate the findings, we analysed differential gene expression data from 12 POP patients. Vaginal fibroblasts from 4 women with POP were used to test the effect of the anti-diabetic drug metformin.

**Outcome measurements and statistical analysis:** The exome chip study used a case-control design to identify single nucleotide variants (SNVs) associated with POP after Bonferroni correction. The molecular landscape was built using the UniProt and PubMed databases to identify functional interactions between the POP candidate genes/proteins. We performed enrichment and upstream regulator analyses of the differentially expressed genes. The effect of metformin in fibroblasts was assessed using one-sample t-test.

**Results and limitations:** We found significant association between POP and SNVs in 54 genes. The proteins encoded by 26 of these genes fit into a molecular landscape, together with 37 other POP candidate molecules and two POP-implicated microRNAs. This landscape is located in and around epithelial cells and fibroblasts of the urogenital tract and harbors four interacting biological processes - epithelial-mesenchymal transition, immune response, modulation of the extracellular matrix, and fibroblast function - that are regulated by sex hormones and TGFB1. Based on the landscape, we predicted and showed that metformin alters gene expression in fibroblasts of POP patients in a beneficial direction. The main limitation of our study is that we have no independent replication of the exome chip results.

**Conclusions:** The integrated molecular landscape of POP that we built provides insights into the biological processes underlying the disease and clues towards novel treatments.

**Patient summary:** We reported the first exome chip study of POP and combined the genes identified in this study with other data from the literature to build a ‘molecular landscape’ of POP. This landscape will advance our understanding of the disease and may lead to novel treatments.

## Introduction

Pelvic organ prolapse (POP) represents a major health care burden in women, with a reported prevalence of up to 40%^1,2^. POP is not life-threatening but its bothersome symptoms have a significant impact on quality of life. Furthermore, POP frequently co-occurs with stress urinary incontinence (SUI), with POP and SUI constituting the two main pelvic floor disorders^3^. The life-time risk for any surgical intervention for POP - with or without SUI - is approximately 20%^4^ and the total costs for POP surgery are substantial^5^. POP has a multifactorial etiology in which both hereditary and environmental factors play a role. Parity, vaginal delivery, age, and high body mass index (BMI) have been identified as environmental risk factors^6^ and could all cause defects of pelvic floor tissues. Familial susceptibility to these defects seems likely, since women with POP more often have family members with POP compared to women without POP^7,8^. Evidence for a genetic background of POP has also emerged from twin studies, with approximately 40% of the POP liability being explained by genetic factors^9,10^. Candidate gene association studies have found a number of single nucleotide polymorphisms (SNPs) to be putatively associated with POP, including genes encoding steroid hormone receptors, collagens type I and III, and multiple matrix metalloproteinases. However, only one SNP, rs1800012 in the *COL1A1* gene, has demonstrated replicable association with POP^11^. In addition, a genome-wide association study (GWAS) identified five SNPs associated with POP that were nominally replicated^12^. As is the case in many other fields of medicine, the translational step from genetic data towards clinically useful information – i.e. diagnostic biomarkers and disease-modifying treatments – still needs to be made in the field of POP. Therefore, in the present study, we have integrated the results from the first exome chip study of POP – which suggested multiple novel candidate genes – with other available genetic and expression data to generate and corroborate a molecular landscape of POP. This landscape provides more detailed and novel insights into the biological processes that, when impaired, could be involved in disease pathogenesis. Furthermore, the POP landscape provides leads for existing drugs that could be repurposed and for novel, disease-modifying treatments of POP that could be developed.

## Methods and Materials

### Exome chip study

#### Participant characteristics

A case-control design was used for the exome chip study. After quality control (QC), the case group consisted of 526 women who presented with symptoms and signs of POP (some of whom had recurrent POP) at the department of Obstetrics and Gynecology of the Radboud university medical center (Radboudumc), Nijmegen, The Netherlands, in January 2007-February 2011. POP was defined as stage II or more, according to the Pelvic Organ Prolapse Quantification (POP-Q) system^13^, i.e. descent of the leading edge of the prolapse of at least 1 cm above the hymenal remnants. POP had already been surgically treated in some women at the time of inclusion. Exclusion criteria were a poor understanding of the Dutch language and/or genetic diseases with an increased risk of POP (e.g., Ehlers-Danlos and Marfan syndromes, and myotonic dystrophy type 1). For POP cases, we collected data on previous surgery, POP-Q stage as well as POP complaints measured with the validated Dutch translation of the Urogenital Distress Inventory (UDI)^14^. In case more than one measurement was available over time, the most severe measurement was used. The UDI records the presence or absence of specific symptoms, and associated bother on a four-point Likert scale. The UDI scores are transformed to a continuous scale, ranging from 0 (no bother) to 100 (greatly bothersome). According to this continuous scale, women without symptoms and women with symptoms but without any bother are scored equally. The exome chip study was approved by the local institutional review board in an amendment of CMO numbers 2007/043 and 2010/071. As the control group, we used the exome chip data available from 960 women from the Nijmegen Biomedical Study (NBS), a general population-based survey conducted by Radboudumc’s Departments for Health Evidence and Department of Laboratory Medicine^15^. Because the controls were sampled from the general population and due to the high prevalence of POP in the population, it is expected that some of the controls had POP as well. For the cases and controls, data on age, parity, and BMI were collected.

For further details of the materials and methods used in this study - including details about QC, population structure and statistical analysis for the exome chip study, and more information about the molecular landscape building, enrichment analysis of expression data from an independent POP cohort, and in vitro experiments - see **Supplementary Materials and Methods**.

## Results

### Exome chip study

We conducted an exome chip study with 526 POP cases and 960 healthy controls. The characteristics of all participants in the exome chip study are presented in **Table 1**. Compared to cases, controls were on average 5 years older and had a higher ratio of postmenopausal females. Since the prevalence of POP increases with age and is probably associated with postmenopausal state – making it likely that some of the controls had POP as well – the results of our study may be underestimating the real genetic differences. We identified 54 statistically significant - i.e., with Bonferroni-corrected P < 0.05, uncorrected P < 1.48E-06 - exonic and (potentially) deleterious single nucleotide variants (SNVs), including 50 missense mutations, 1 splice site mutation and 1 stop-gain mutation. The list of these SNVs and the genes they affect is provided in **Table 2**. Interestingly, of the 54 SNVs for which a Combined Annotation-Dependent Depletion (CADD) score was available – a single pathogenicity score that combines 63 distinct computational prediction methods^17^, including the Grantham score for missense mutations^18^ – 18 had a CADD score > 20 and 12 had a CADD score > 10 but < 20 (**Table 2**). These SNVs are predicted to be among the 1 and 10% most deleterious of all possible substitutions in the human genome, respectively^17,19^. This implies that a considerable proportion of the SNVs that we identified have a damaging effect on protein function.

**Table 1.** Characteristics of POP cases and controls that were analyzed in our exome chip study.

|  |  |  |  |
| --- | --- | --- | --- |
| Characteristic | Cases (n=526) * | Controls (n=960) * | P value |
| Age <sup>a</sup> (years) | 51.3 ± 13.5 [21-87; 522] | 56.2 ± 10.8 [27-75; 960] | < 0.01 <sup>e</sup> |
| Parity | 2.4 ± 1.1 [0-9; 515] | 2.3 ± 1.1 [0-10; 791] | 0.47 <sup>e</sup> |
| BMI (kg/m<sup>2</sup>) | 25.6 ± 3.9 [15-42; 393] | 25.4 ± 4.3 [17.6-50.1; 937] | 0.29 <sup>e</sup> |
| Previous POP surgery<br>0<br>1<br>≥2 | 277 (52.8; 525)<br>150 (28.6; 525)<br>98 (18.7; 525) | unknown | NA |
| POP-related complaints <sup>b</sup> | 48.9 ± 32.3 [0-100; 323] | unknown | NA |
| POP-Q stage <sup>c</sup><br>Stage 0<br>Stage I<br>Stage II<br>Stage III<br>Stage IV | 2 (0.4; 502)<br>12 (2.4; 502)<br>217 (43.2; 502)<br>248 (49.4; 502)<br>23 (4.6; 502) | unknown | NA |
| Postmenopausal <sup>d</sup> | 278 (73; 380) | 771 (81; 955) | < 0.01 <sup>f</sup> |

\* Data are presented as mean ± standard deviation [range; n] in case of continuously distributed data and as absolute numbers (percentages; n) in case of categorical data.
<sup>a</sup> Age at diagnosis for cases, age at time of survey/sampling for controls.
<sup>b</sup> Measured with and reported as 'total score' from the validated Dutch translation of the Urogenital Distress Inventory (UDI). In case of repetitive measurements over time, the worst UDI score available in the medical file was used.
<sup>c</sup> Because the genetic background does not change over time, women were included in the exome chip study as a POP case at any time after diagnosis. This could also be after successful reconstructive surgery, which explains the lower stages in some POP cases (who had already been surgically treated). In all POP cases, the worst stage available in the medical file was used.
<sup>d</sup> This was assessed through the NBS participants' answer to the question: 'Did you ever experience menopausal symptoms (e.g. hot flushes)', with possible 'yes', 'no' or 'not applicable' (I am not in the menopause yet).
<sup>e</sup> P value calculated with independent samples t-test.
<sup>f</sup> P value calculated with chi-square test.
Abbreviations: BMI, body mass index; NA, not applicable; POP, pelvic organ prolapse; POP-Q, Pelvic Organ Prolapse Quantification system.

**Table 2.** Top 54 SNVs identified through our exome chip study. For each SNV, the minor allele frequencies in the POP cases and controls (MAF-ca and MAF-co), the P value of association with POP and the Bonferroni-corrected P value are provided. All SNPs with Bonferroni-corrected P < 0.05 are listed. In addition, the affected gene, SNV type, nucleotide change and amino acid change are indicated. If applicable and available, the Grantham and CADD scores – which are both predictive measures of pathogenicity – for each SNV are given. Further, the 26 genes encoding proteins within the molecular landscape of POP (**Figure 1**) are indicated in bold.

| SNV | MAF-ca <sup>a</sup> | MAF-co <sup>a</sup> | P | Corr P | Gene | Type | Nucleotide change | Amino acid change | GS <sup>b</sup> | CADD <sup>c</sup> |
| --- | --- | --- | --- | --- | --- | --- | --- | --- | --- | --- |
| rs138799629 | 0 | 0,13650 | 6,25E-36 | 2,11E-31 | *ANKRD49* <sup>d</sup> | missense | G>A | p.Val46Ile. | 29 | 14.58 |
| rs115986826 | 0,08175 | 0,00208 | 7,41E-34 | 2,51E-29 | **NEB** | missense | T>G | p.Asp1329Ala | 126 | 24.70 |
| rs138533962 | 0,00380 | 0,13490 | 2,96E-33 | 1,00E-28 | **STYK1** | missense | G>A | p.Arg379Cys | 180 | 25.2 |
| rs149745899 | 0,00285 | 0,11880 | 2,17E-29 | 7,35E-25 | **SCUBE2** | missense | G>A | p.Ser481Phe. | 155 | 18.81 |
| rs140794579 | 0,03422 | 0,17660 | 9,56E-29 | 3,24E-24 | **ACN9** | missense | A>C | p.Gln97Pro | 76 | 26.4 |
| rs199737061 | 0,01616 | 0,13700 | 5,54E-27 | 1,88E-22 | *ZNF700* <sup>e</sup> | missense | A>C | p.His685/703Pro | 77 | 0.001 |
| rs138091786 | 0,00856 | 0,12140 | 8,10E-27 | 2,74E-22 | *LRRC34* | missense | A>G | p.Val308/324/337/369Ala. | 64 | 14.38 |
| rs114675387 | 0,00382 | 0,10890 | 4,07E-26 | 1,38E-21 | *MUC17* | missense | A>G | p.Thr2611Ala | 58 | 0.002 |
| rs61748812 | 0,05133 | 0,17760 | 3,62E-22 | 1,22E-17 | **NACA2** | missense | C>A | p.Val9Phe | 50 | 17 |
| rs150552771 | 0 | 0,07865 | 1,25E-20 | 4,25E-16 | *LPGAT1* <sup>f</sup> | missense | T>C | p.Lys200Glu. | 56 | 19.09 |
| rs151097728 | 0,00095 | 0,08021 | 1,96E-20 | 6,64E-16 | **TFAM** | missense | C>G | p.Gln82/100Glu | 29 | 0.013 |
| rs11828907 | 0,04373 | 0,00052 | 1,34E-19 | 4,54E-15 | **AHNAK** | missense | T>C | p.Asp4304Gly | 94 | 22.60 |
| rs144392720 | 0,04183 | 0 | 1,51E-19 | 5,13E-15 | **TNFRSF10C** | missense | A>G | p.Gln58Arg | 43 | 3.575 |
| rs200940950 | 0,00190 | 0,07500 | 1,61E-18 | 5,46E-14 | **METAP1** | splice site | C>T | - | NA | 5.733 |
| rs79324787 | 0 | 0,06719 | 9,80E-18 | 3,32E-13 | *KIF27* | missense | T>C | p.Ile890/921/987Val | 29 | 2.241 |
| rs142757633 | 0 | 0,06719 | 1,05E-17 | 3,57E-13 | **YWHAB** | missense | A>C | p.Lys77Gln | 53 | 17.12 |
| rs78707986 | 0,00190 | 0,07031 | 2,63E-17 | 8,91E-13 | *GXYLT1* | missense | A>C | p.His117/148Gln | 24 | 22.30 |
| rs141705162 | 0,00095 | 0,06667 | 6,02E-17 | 2,04E-12 | **FASTKD1** | missense | G>A | p.Ala320/343Val | 64 | 19.38 |
| rs76146040 | 0,00570 | 0,07604 | 1,17E-16 | 3,96E-12 | *OR5H14* | missense | G>A | p.Met234Ile | 10 | 0.051 |
| rs34429135 | 0,03707 | 0,13120 | 1,58E-16 | 5,34E-12 | **LAIR2** | missense | T>A | p.Phe115Tyr. | 22 | 0.001 |
| rs142631876 | 0 | 0,06094 | 4,05E-16 | 1,37E-11 | **RRM2** | missense | A>G | p.Arg342/402Gly | 125 | 19.83 |

Table 2 – continued.
| SNV | MAF-ca <sup>a</sup> | MAF-co <sup>a</sup> | P | Corr P | Gene | Type | Nucleotide change | Amino acid change | GS <sup>b</sup> | CADD <sup>c</sup> |
| --- | --- | --- | --- | --- | --- | --- | --- | --- | --- | --- |
| rs145531717 | 0,00095 | 0,06302 | 4,30E-16 | 1,45E-11 | **ATP12A** | missense | C>A | p.Arg490/496Ser | 110 | 23.80 |
| rs151149890 | 0 | 0,05885 | 1,10E-15 | 3,71E-11 | *LRRC37A3* | missense | G>A | p.Thr570/631/711/1593Met | 81 | 0.003 |
| rs200909708 | 0,00570 | 0,07240 | 1,12E-15 | 3,79E-11 | **AKR1C1** <sup>g</sup> | missense | G>A | p.Val118/151/153Met | 21 | 0.133 |
| rs146515657 | 0,05038 | 0,00573 | 1,72E-15 | 5,83E-11 | **USP4** | missense | T>C | p.Asn389/603/650Ser | 46 | 3.361 |
| rs190805303 | 0,36410 | 0,50780 | 5,08E-14 | 1,72E-09 | *CCDC90B* | missense | T>C | p.Thr90/118/182/191Ala | 58 | 0.131 |
| rs144738877 | 0 | 0,05312 | 3,07E-14 | 1,04E-09 | *IFT80* | missense | C>T | p.Arg44/116/253/424His | 29 | 27.90 |
| rs4362173 | 0,03517 | 0,11090 | 1,16E-12 | 3,92E-08 | *OR52E6* | missense | T>C | p.Ile39/43Val | 29 | 0.001 |
| rs149907018 | 0 | 0,04740 | 7,55E-13 | 2,55E-08 | *SCN4A* | missense | T>C | p.Ile430Val | 29 | 5.153 |
| rs138362833 | 0,40210 | 0,53650 | 2,93E-12 | 9,91E-08 | *PRSS1* | missense | A>C | p.Lys237/251Asn | 94 | 9.898 |
| rs114501427 | 0 | 0,04531 | 2,77E-12 | 9,36E-08 | **SPOPL** | missense | G>A | p.Asp349Asn | 23 | 20.90 |
| rs2302212 | 0,05989 | 0,13330 | 6,23E-10 | 2,11E-05 | *NCAPG* | missense | T>C | p.Met231Thr | 81 | 15 |
| rs144426665 | 0 | 0,03750 | 2,70E-10 | 9,15E-06 | *OR5B17* | missense | C>A | p.Val124Leu | 32 | 25.50 |
| rs115910467 | 0,00286 | 0,04271 | 5,77E-10 | 1,95E-05 | **GGCT** | missense | C>T | p.Arg108/160His | 29 | 17.43 |
| rs79820636 | 0,40820 | 0,52190 | 2,94E-09 | 9,96E-05 | *PCMTD1* | missense | C>T | p.Arg172/248His | 29 | 34 |
| rs4987208 | 0,04485 | 0,01042 | 1,53E-09 | 5,18E-05 | **RAD52** <sup>h</sup> | stop-gain | A>C | p.Tyr415Ter | NA | 35 |
| rs201483250 | 0,00761 | 0,05052 | 2,04E-09 | 6,90E-05 | **LILRA1** | missense | C>T | p.Arg179/196Trp | 101 | 10.48 |
| rs201702426 | 0,01616 | 0,06458 | 2,43E-09 | 8,23E-05 | **AKR1D1** | missense | T>C | p.Ile63/119Thr | 89 | 25.90 |
| rs9871143 | 0,56840 | 0,46090 | 2,14E-08 | 7,23E-04 | *OR5H6* | missense | G>A | p.Asp285Asn | 23 | 12 |
| rs4252119 | 0 | 0,03281 | 3,28E-09 | 1,11E-04 | **PLG** | missense | C>T | p.Arg408Trp | 101 | 0.880 |
| rs200020201 | 0,00095 | 0,03490 | 3,33E-09 | 1,13E-04 | *NKD1* | missense | A>G | p.Met130Val | 21 | 0.089 |
| rs200454083 | 0,02376 | 0,07708 | 3,63E-09 | 1,23E-04 | **ANKLE2** | missense | G>C | p.Pro175/758/820Ala | 27 | 0.001 |
| rs17115182 | 0 | 0,03125 | 7,78E-09 | 2,63E-04 | *KRR1* | missense | G>A | p.Pro43Ser | 74 | 24.40 |
| rs201566504 | 0 | 0,03021 | 1,75E-08 | 5,91E-04 | *ZFP14* <sup>i</sup> | missense | C>T | p.Val322Ile | 29 | 23.00 |
| rs11229158 | 0,00095 | 0,02552 | 5,43E-07 | 1,84E-02 | *OR4A16* | missense | C>A | p.Leu188Ile | 5 | 8 |

Table 2 – continued.
| SNV | MAF-ca <sup>a</sup> | MAF-co <sup>a</sup> | P | Corr P | Gene | Type | Nucleotide change | Amino acid change | GS <sup>b</sup> | CADD <sup>c</sup> |
| --- | --- | --- | --- | --- | --- | --- | --- | --- | --- | --- |
| rs141135045 | 0 | 0,02865 | 3,67E-08 | 1,24E-03 | *RFESD* | missense | C>G | p.Pro81/134Arg | 103 | 31.00 |
| rs117389731 | 0,00951 | 0,04740 | 5,93E-08 | 2,01E-03 | *ZNF780B* | missense | G>A | p.Thr542/690Ile | 89 | 0.086 |
| rs1051949 | 0 | 0,02760 | 6,81E-08 | 2,31E-03 | ***MRPL35*** | missense | G>A | p.Arg29His | 29 | 24.00 |
| rs200482299 | 0,00095 | 0,02917 | 9,59E-08 | 3,25E-03 | ***ARHGEF19*** | missense | C>T | p.Ala157/163/474Thr | 58 | 24.20 |
| rs200171449 | 0 | 0,02500 | 2,48E-07 | 8,40E-03 | *TAS2R46* | missense | T>G | p.Ser170Arg | 110 | 0.629 |
| rs188761759 | 0,00856 | 0,04115 | 6,81E-07 | 2,30E-02 | ***EMILIN1*** | missense | G>T | p.Ala391Ser | 99 | 0.405 |
| rs74951519 | 0,36410 | 0,50780 | 5,08E-14 | 1,72E-09 | *OR7C2* | missense | A>G | p.Lys294Arg | 26 | 0.002 |
| rs144462785 | 0,00095 | 0,02552 | 6,93E-07 | 2,34E-02 | *PCDHB12* | missense | T>G | p.Leu252Arg | 102 | 0.036 |
| rs17213351 | 0,00951 | 0,04219 | 7,62E-07 | 2,58E-02 | *HLA-DQA2* | missense | G>A | p.Ala222Thr | 58 | 22.40 |

Abbreviations: NA, not applicable; POP, pelvic organ prolapse; SNV(s), single nucleotide variant(s)
<sup>a</sup>. A MAF of '0' indicates that none of the POP cases or controls were found to carry the minor allele of the SNV; <sup>b</sup>. the Grantham score (GS) for missense mutation-type SNVs ranges from 0 to 215 and a higher score indicates a stronger negative effect of the amino acid change on protein structure and function<sup>18</sup>; <sup>c</sup>. the Combined Annotation-Dependent Depletion (CADD) tool combines 63 distinct computational prediction methods – including the GS for missense mutations – and calculates a single pathogenicity score<sup>17</sup>. SNVs with CADD scores > 20 and > 10 indicate that these are predicted to be among the 1 and 10% most deleterious of all possible substitutions in the human genome, respectively<sup>17,19</sup>. Within our set of differentially expressed genes in an independent POP cohort (see Methods), the expression of six of the genes from the exome chip was different: the expression of the expression of *ANKRD49* was downregulated (FC=-1,19; corrected P=7.08E-03)<sup>d</sup>, the expression of *ZNF700* was downregulated (FC=-1,13; corrected P=2.27E-02)<sup>e</sup>, the expression of *LPGAT1* was upregulated (FC=1,19; corrected P=1.70E-02)<sup>f</sup>, *AK1RC1* was upregulated (FC=1,55; corrected P=7.01E-03)<sup>g</sup>, the expression of *RAD52* was downregulated (FC=-1,16; corrected P=3.77E-02)<sup>h</sup>, and the expression of *ZFP14* was downregulated (FC=-1,32; corrected P=3.94E-02)<sup>i</sup>.

### Molecular Landscape of POP

A molecular landscape was built in which 26 of the 54 proteins encoded by the genes from the exome chip study (**Table 2**) interact functionally, together with 37 other POP candidates as well as 2 POP-implicated microRNAs (**Supplementary Table 1**). The molecular landscape of POP is shown in **Figure 1** and includes 11 proteins/molecules – indicated in white – that have not been directly linked to POP (yet) but show important functional interactions within the landscape. The molecular POP landscape is located in epithelial cells, fibroblasts and surrounding extracellular matrix (ECM) of the female urogenital tract. Four main biological processes operate within the landscape, i.e. epithelial-mesenchymal transition (EMT), immune response activation, modulation of the ECM, and fibroblast survival and apoptosis. These processes interact with each other and are regulated through signaling cascades involving female hormones and by the cytokine transforming growth factor beta 1 (TGFB1). In the legend of **Figure 1**, a succinct description of these four processes is given and in the **Supplementary Information**, a detailed molecular signaling and interactions is provided.

**Figure 1.**
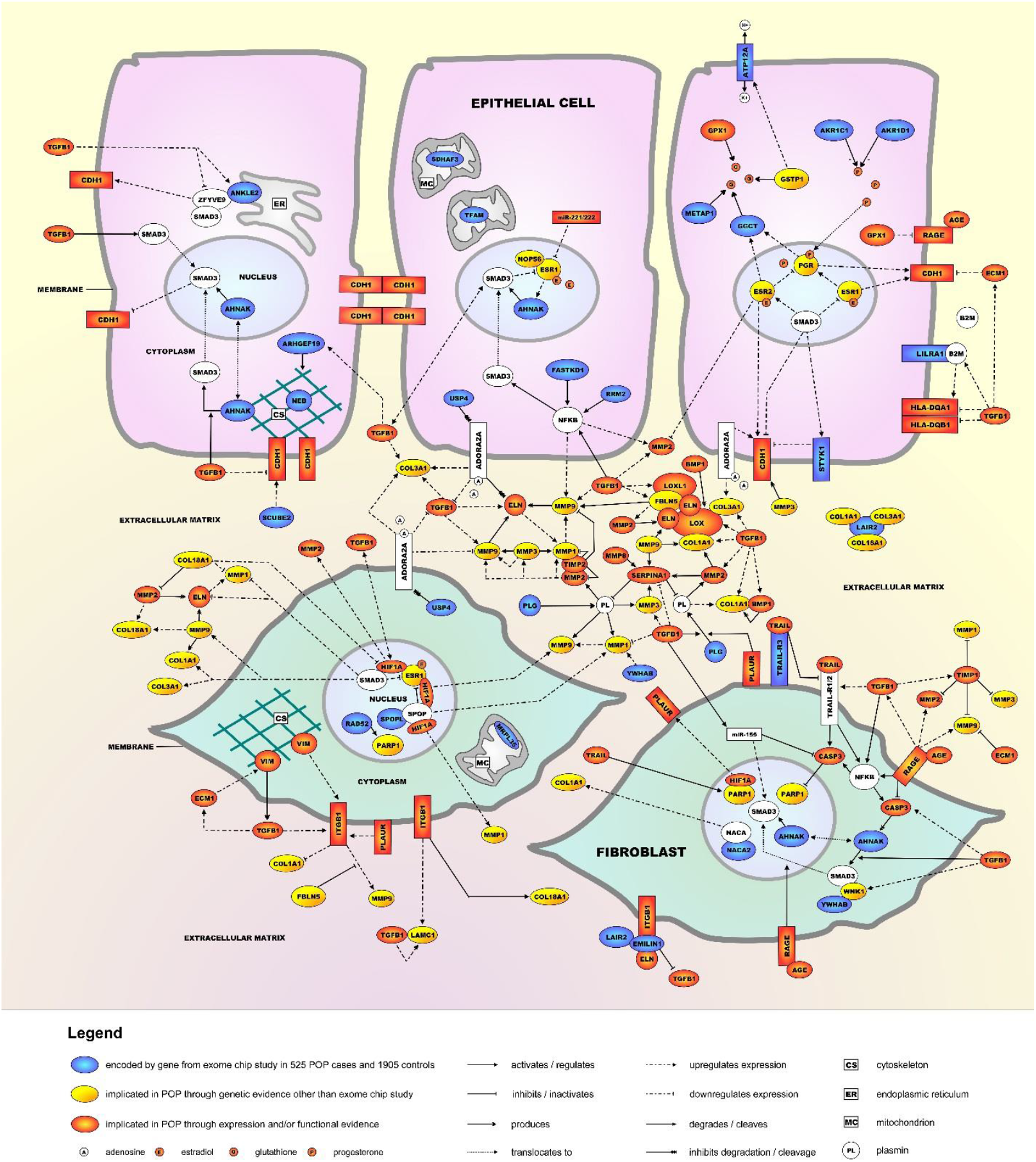
Molecular landscape of POP. The POP landscape is located in and around epithelial cells and fibroblasts of the female urogenital tract. Four main biological processes operate in the landscape: **(1)** The first and most important process in the landscape is EMT. Under normal conditions, epithelial cells are connected to their environment through cell-cell interactions as well as cell-ECM interactions. Among the cell-cell interactions are epithelial cadherin (E-cadherin/CDH1)-based adherens junctions that stabilize epithelial cell-cell adhesion. EMT is characterized by a ‘cadherin switch’, which is initiated when epithelial cells start producing less CDH1 and more of the mesenchymal marker N-cadherin (CDH2) (not shown). This results in loss of epithelial cell-cell adhesion and the epithelial cells gradually transforming into mesenchymal cells which then further differentiate into fibroblasts. EMT involves the key transcription factor SMAD3 and is negatively regulated by three sex hormone-bound receptors – the estrogen receptors ESR1 and ESR2 and the progesterone receptor (PGR) – that each upregulate CDH1 expression and hence prevent EMT. Conversely, TGFB1 induces EMT as it downregulates CDH1 expression. **(2)** Proteins of the major histocompatibility complex II (MHC II) – i.e. HLA-DQA1 and HLA-DQB1 – are expressed in epithelial cells of the female reproductive tract, where they are involved in regulating the activity of the immune response through presenting foreign antigens to circulating T lymphocytes (not shown). TGFB1 regulates the expression of these two HLA proteins, in part through upregulating the expression of beta-2-microglobulin (B2M), an EMT-inducing extracellular protein that mediates antigen presentation **(3)** Further, the ECM provides support to epithelial cells and fibroblasts and modulation of the ECM is essential for many pathophysiological processes such as tissue growth, wound healing, and fibrosis. The ECM is composed of different molecules, including elastin (ELN) and collagen (COL) fibers as well as proteins that crosslink or regulate the degradation of ELN and COL (e.g. LOX and LOXL1), various matrix metalloproteinases (MMPs) and their regulators TIMP1 and TIMP2. Other important ECM proteins are ECM1, FBLN5, LAIR2 and LAMC1. TGFB1 is involved in ECM remodeling through regulating the expression of most ECM proteins in the landscape. **(4)** Since fibroblasts are responsible for the synthesis and secretion of the main ECM components through SMAD3 – which, as indicated above, also plays a major role in EMT – fibroblast survival and apoptosis (and hence proper functioning) is another important landscape process. TRAIL, a cytokine of the tumor necrosis factor (TNF) family, regulates fibroblast survival and apoptosis through its receptors and signaling involving pro-apoptotic proteins such as NFKB and CASP3. As with the three other main processes, TGFB1 is a key regulator, e.g. through regulating the expression of TRAIL-R1 and CASP3, and activating NFKB. In addition, extracellular AGE (or advanced glycation end product) molecules binding to their receptor RAGE results in NFKB activation, CASP3 inhibition and translocation of SMAD3 to the nucleus, which results in the AGE-RAGE complex stimulating the apoptosis of fibroblasts. Lastly, nuclear proteins such as HIF1A and PARP1 are also important regulators of fibroblast survival and apoptosis.

### Enrichment analyses of differential expression data from an independent POP patient cohort

The upstream regulator analysis of the 618 genes that were differentially expressed in an independent POP patient cohort at a corrected P < 0,01 (**Supplementary Tables 2 and 3**) yielded five regulators that are also present in the POP landscape: TGFB1, SMAD3, PGR and ESR2 are predicted to be activated while COL18A1 is predicted to be inhibited (**Table 3a**). Moreover, the targets of TGFB1 are most significantly enriched within the set of 618 genes. We identified five functional gene categories that are present in the POP landscape and are enriched within the 618 differentially expressed genes: three categories relating to (epithelial) cell death – i.e., ‘apoptosis’, ‘necrosis of epithelial tissue’ and ‘cell death of epithelial cells’ – are predicted to be inhibited while two categories relating to (fibroblast) survival – i.e., ‘cell survival’ and ‘differentiation of fibroblasts’ – are predicted to be activated (**Table 3b**). We also identified five genes that encode proteins within the POP landscape *and* are differentially expressed between POP and non-POP tissues in an independent POP patient cohort, i.e. *AKR1C1, PLAUR* and *VIM* are upregulated while *FBLN5* and *RAD52* are downregulated (**Supplementary Table 4**).

**Table 3a.**
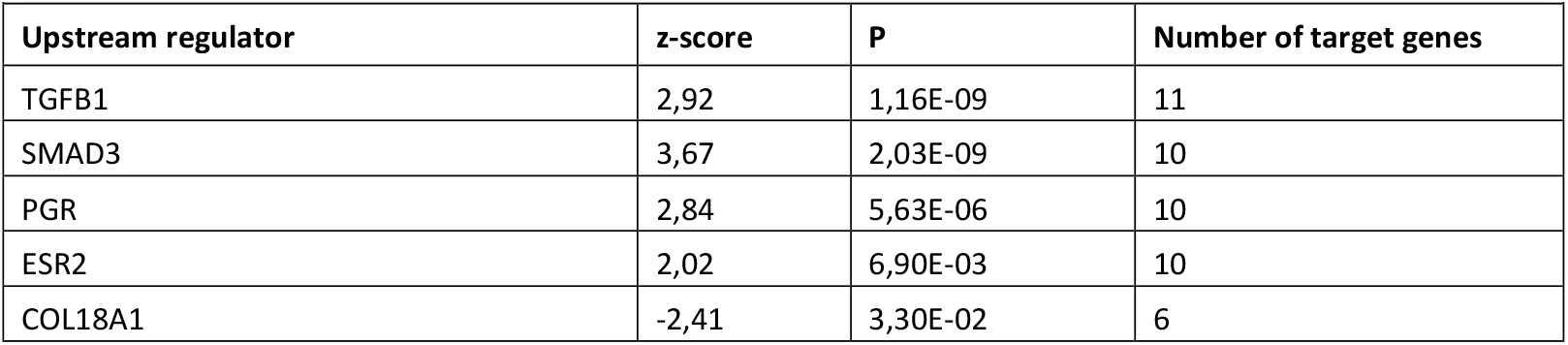
Upstream regulator analysis of the 618 mRNAs that were differentially expressed (corrected P < 0,01) between POP and non-POP tissues in 12 POP patients (see Supplementary Materials and Methods). The 5 upstream regulators that are present in the POP landscape are listed with their z-score ≥ 2,00 or ≤ -2,00, indicating that they are predicted to be activated or inhibited, respectively. For each regulator, the P-value – referring to how enriched the targets of each upstream regulator are in the differentially expressed mRNAs – and the number of target genes are also listed.

**Table 3b.** Functional gene categories that are present in the POP landscape and enriched within the 618 mRNAs that were differentially expressed (corrected P < 0,01) between POP and non-POP tissues in 12 POP patients (see Supplementary Materials and Methods). The 5 categories are listed with their z-score ≥ 2,00 or ≤ -2,00, indicating that they are predicted to be activated or inhibited, respectively. For each category, the BH-corrected P-value of enrichment and the number of genes in the category are also listed.

| Gene category | z-score | BH-corrected P | Number of genes |
| --- | --- | --- | --- |
| Apoptosis <sup>a</sup> | -2,47 | 1,23E-12 | 185 |
| Cell survival <sup>a</sup> | 2,34 | 2,06E-07 | 108 |
| Necrosis of epithelial tissue <sup>a</sup> | -2,52 | 7,92E-05 | 49 |
| Differentiation of fibroblasts <sup>b</sup> | 2,55 | 7,63E-04 | 11 |
| Cell death of epithelial cells <sup>a</sup> | -2,51 | 2,01E-03 | 38 |

Abbreviations: POP, pelvic organ prolapse; BH, Benjamini Hochberg.
<sup>a</sup> This gene category falls under the broader 'cell death and survival' category.
<sup>b</sup> This gene category falls under the broader 'connective system development and function' category.

### Metformin downregulates the expression of TGFB1 target genes in vaginal fibroblasts from POP patients

As TGFB1 is the ‘master regulator’ of the POP landscape and its activation results in many landscape processes that are involved in POP, we tried to confirm metformin – an oral anti-diabetic drug that counteracts TGFB1 signaling – as a drug target ‘lead’ for POP by testing its effect on the expression of selected genes from the landscape. Since metformin has been shown to suppress TGFB1 responses specifically in fibroblasts cell lines^20^, we used human fibroblasts from patients to see if the drug could have the same beneficial effect in POP. As shown in **Figure 2**, we found that metformin affects the TGFB1-induced expression of three ECM proteins in these fibroblasts: *COL3A1* (P=0,0136) and *ELN* (P=0,039) were significantly downregulated, while there was a tendency towards lower expression of *COL1A1* (P=0,0961).

**Figure 2.**
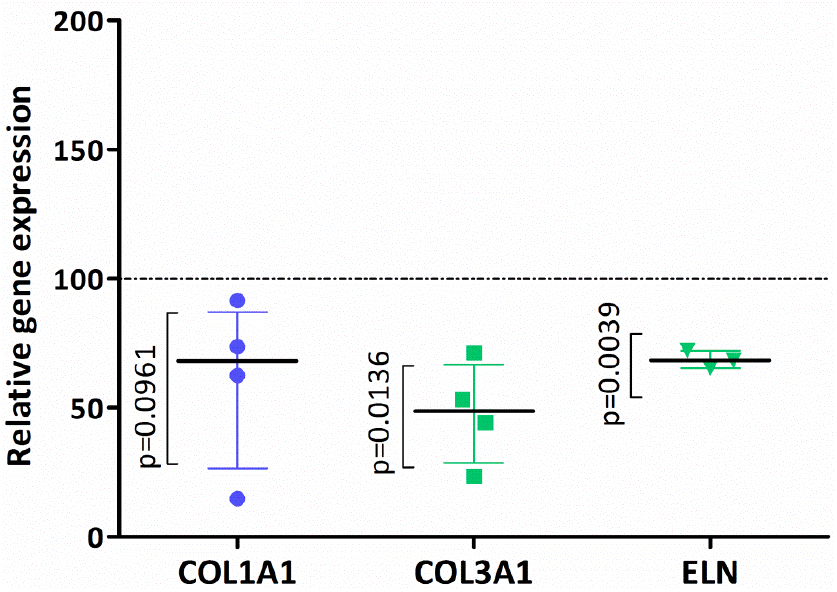
Metformin downregulates TGFB1-induced gene expression in fibroblasts from women with POP. Cells derived from prolapsed tissues from premenopausal women (n=4) were stimulated for 24 hours with TGFB1 [0.1ng/ml] and then challenged with Metformin [2mM] for another 24 hours. Real time PCR was used to measure three TGFB1-regulated genes from the landscape, i.e. the collagen genes *COL1A1* and *COL3A1* and *elastin (ELN)* and the two house-keeping genes YWHAZ and HPRT1. The results are the relative gene expression and are shown as a percentage of TGFB1-stimulated cells. The dotted lines are 100% corresponding to the gene expression of the cells treated with TGFB1 [0.1ng/ml]. One outlier was identified using Grubbs’ test^58^ and removed from the ELN gene expression data set (n=3). P-values were calculated using one-sample t-test compared to 100.

## Discussion

We integrated the available genetic and expression data with our results from the first exome chip study of POP into a protein interaction landscape of the disease. This molecular landscape contains the proteins encoded by 26 of the 54 genes affected by SNVs from the exome chip study, as well as 37 other POP candidate molecules and two POP-implicated microRNAs. The POP landscape reveals four main biological processes that operate in epithelial cells, fibroblasts and surrounding ECM of the female urogenital tract: EMT, immune response activation, ECM modulation, and fibroblast survival and apoptosis. Of note, none of the “classical” POP candidate genes – identified through candidate gene association studies and mainly encoding ECM proteins – or the genes from the only thus far conducted GWAS of POP (including the landscape genes *COL18A1* and *NOP56*) contained significant SNVs in our study. Moreover, our landscape does not assume or represent a “sequence of events” i.e., a number of biological processes and signaling cascades that occur in a spatially and temporally distinct order. Instead, a deficit in any or a combination of the four main landscape processes or their constituting signaling cascades, possibly aggravated through environmental risk factors and caused by variants in one or more genes, can lead to POP in various degrees of severity. This is reflected by the clinical reality of POP as a disease with a multifactorial etiology, a complex and variable symptom presentation and variable treatment response.

Multiple signaling cascades in the landscape are regulated by female sex hormones (i.e. estrogen in its active form estradiol, and progesterone). These hormones indirectly affect oxidative stress levels in epithelial cells and fibroblasts of the urogenital tract, as progesterone-activated PGR^21^ and estradiol-activated ESR2^22^ upregulate the expression of the enzyme GGCT. Together with three other enzymes from the landscape – GPX1, GSTP1 and METAP1 – GGCT is involved in the metabolism of glutathione^23^, an important antioxidant that prevents oxidative stress-induced cellular damage by reactive oxygen species (ROS)^24^. Interestingly, ROS production by fibroblasts of women without POP was found to be increased through mechanical stress^25^ while oxidative stress biomarkers were also increased in the uterosacral ligaments of POP patients^26^. ROS can also directly induce EMT^27^, corroborating the role of oxidative stress in POP. Although sex hormones influence many landscape cascades, their role in POP etiology is not completely understood. Increasing age and postmenopausal state are risk factors for POP^6^ and the hypoestrogenic state after the menopause could be a factor in the onset of POP. Both lower serum levels of estradiol^28-30^ and lower serum and urine levels of progesterone (metabolites)^28,31^ have been associated with an increased risk of POP. Hence, hormone replacement therapy (HRT) in postmenopausal women could have a favorable effect on the disease progression or even prevent the disease from developing. However, studies on the effect of HRT in POP patients show conflicting results, as negative^32^, protective^33^ or no effects^2,34,35^ have been reported. Therefore, we suggest that future studies on the effect of HRT on POP take into consideration confounding factors such as endogenous sex hormone levels, menopause duration or whether POP patients have less functioning hormone receptors due to genetic variation.

The cytokine TGFB1 represents the ‘master regulator’ as, when activated, it affects and modulates all four biological processes in the landscape. Notably, (activated) TGFB1 is involved in ECM remodeling and regulating fibroblast function. Moreover, TGFB1 itself has an important role in POP etiology, and TGFB1 is both differentially expressed^30,36^ and could have a functional effect – e.g. through affecting fibroblast proliferation^37^ – in POP patients.

We have been able to corroborate our findings by analyzing differential gene expression data from an independent POP cohort. Our analysis of these data revealed that four ‘upstream regulators’ – the most significant regulator TGFB1 and three other proteins that are important in EMT: SMAD3 and the sex hormone receptors PGR and ESR2 – are predicted to be activated in POP tissue while COL18A1, a collagen with multiple roles in ECM modulation, is predicted to be inhibited. These results are in line with the landscape in which EMT and ECM modulation are two of the four main processes. Moreover, five genes encoding proteins in the landscape – including *AKR1C1* and *RAD52*, two genes from the exome chip study – are differentially expressed in the independent cohort, providing a direct validation of these genes as POP candidates. Further, our findings from the enrichment analysis of functional gene categories indicate that both epithelial cells and fibroblasts are affected in POP.

As the ‘master regulator’ of the landscape, TGFB1 would be an interesting target for developing novel treatments of POP. More specifically, drugs that lower TGFB1 activity could have beneficial effects. However, as a result of its diverse effects on multiple signaling cascades in the body, drugs (negatively) targeting the expression or function of the genes encoding TGFB1 or its receptors are likely to be associated with multiple side effects. Therefore, it would be better to test drugs that selectively target individual signaling pathways downstream of TGFB1. Searching the literature, we have found a number of such drugs.

First, halofuginone inhibits the phosphorylation and subsequent activation of SMAD3 after TGFB1 stimulation^38,39^. Moreover, in various cell and animal models – including the rat uterus – halofuginone inhibits the synthesis of type 1 collagens, such as the important landscape protein *COL1A1*^40,41^. Clinical trials in humans have also indicated that oral or topical application – as a cream – of halofuginone is safe, well-tolerated and has a beneficial effect – with minimal side effects – on skin diseases^42,43^, making it a good candidate to further evaluate as ‘repurposed’ drug for the prevention and/or treatment of POP. Another ‘drug’ that inhibits TGFB1 signaling and could be tested for POP is hesperitin, a natural compound from citrus fruits that has been studied for a possible beneficial effect in multiple diseases due to its anti-oxidant and anti-inflammatory properties^44^. Like halofuginone, hesperitin decreases TGFB1 signaling through inhibiting SMAD3 phosphorylation and activation^45^ and also directly inhibits EMT in rodent kidneys^46^. Further, simvastatin, a commonly used oral drug to reduce cholesterol levels, inhibits TGFB1-dependent EMT and ROS production of kidney cells^47^ while it also specifically inhibits TGFB1-induced collagen type I production by and SMAD3 phosphorylation in fibroblasts^48^.

Lastly and importantly, a novel and (partial) TGFB1-modulating treatment for POP may be metformin, an oral anti-diabetic drug that affects multiple genes/proteins and processes in the landscape. First, metformin reduces the expression of TGFB1^49,50^ and it decreases TGFB1-promoted loss of CDH1^51,52^, which results in less EMT. Metformin also regulates female sex hormone signaling as it suppresses the expression of miR-221^53,54^ and miR-222^53^, two POP-implicated microRNAs that downregulate ESR1 expression. Metformin inhibits TGFB1-induced synthesis of type 1 collagen by fibroblasts through suppressing SMAD3 phosphorylation^55-57^. Therefore and since metformin specifically suppresses TGFB1 responses in fibroblast cell lines^20^, we used fibroblasts from patients to assess if the drug could affect TGFB1 signaling in a direction that (presumably) has a beneficial effect on POP. We indeed show that, through downregulating the expression of the key ECM (and landscape) proteins *COL1A1*, COL3A1 and ELN, metformin counteracts molecular mechanisms downstream of activated TGFB1 that negatively affect POP, as we predicted from the landscape. However, although these in vitro findings add evidential weight to metformin as a novel POP treatment, further studies are needed to fully develop it as a repurposed drug.

The current results should be viewed in light of some strengths and limitations. The main limitations of our study are that we have no independent replication of the exome chip results and we have only initial in vitro findings for the beneficial effect of one drug that could be repurposed for POP treatment. However, our findings are strengthened by the enrichment analyses of differential expression data from an independent POP cohort. In addition, our findings for metformin could serve as the starting point for further studies.

In conclusion, the molecular landscape that we built based on the results from our exome chip study and available literature data and that we could corroborate through analyzing gene expression data from an independent cohort provides novel insights into the genetic etiology of POP. Furthermore, the landscape provides interesting leads for existing drugs that could be developed as repurposed compounds in selected groups of POP patients.

## Data Availability

The authors confirm that the data supporting the findings of this study are available within the article and its supplementary material. Any extra information is available from the corresponding author, upon reasonable request.

## Acknowledgments

We would like to thank all subjects – both patients and healthy controls – for their active participation in the exome chip study. We thank Mark E. Vierhout, emeritus professor in urogynaecology at Radboud university medical center, Nijmegen, The Netherlands, as well as Leon van Kempen, COO and Scientific Director of the Molecular Pathology Center at Jewish General Hospital, QC, Canada and Jeroen Dijkstra, Lead Quality Assurance Officer and Research Technician at Radboud university medical center, since they all had an important role in candidate gene association studies of POP, which has eventually resulted in this paper. Furthermore, we would like to thank the principal investigators of the Nijmegen Biomedical Study (NBS), i.e. the co-author LAK as well as B. Franke, A.L.M. Verbeek and D.W. Swinkels.

